# Intraoperative Hypotension and Risk of Cuff Blood Pressure Inaccuracy

**DOI:** 10.64898/2026.07.20.26358528

**Authors:** Sohee Kwon, Sungsoo Kim, Daniel Cole, Alan C. Bovik, Edward Giovannucci, Maxime Cannesson

## Abstract

**Background:** Intraoperative hypotension is associated with cardiovascular complications and mortality, making accurate blood pressure monitoring essential. However, the accuracy of noninvasive oscillometric cuff blood pressure (Cuff BP) during hypotension is uncertain. We examined the association between intraoperative hypotension and Cuff BP inaccuracy.

**Methods:** A single-center retrospective cohort included 22,812 adults undergoing noncardiac surgery from April 2013 through November 2023, with 159,782 simultaneous Cuff BP and invasive arterial blood pressure (Arterial BP) pairs. Pairs with arterial systolic BP >120 mmHg or diastolic BP >80 mm Hg were excluded to focus on hypotensive range. Hypotension was defined as arterial mean arterial pressure (MAP) <65 mmHg and categorized as mild (55 to <65), moderate (45 to <55), or severe (35 to <45 mmHg). Cuff BP inaccuracy was defined as an absolute MAP difference >10 mmHg from Arterial BP. Multivariable logistic regression estimated adjusted odds ratios (ORs) and 95% CIs.

**Results:** Compared with normal MAP (≥65 mmHg), hypotension was associated with greater odds of Cuff BP inaccuracy (adjusted OR, 1.45 [95% CI, 1.41–1.49]). Adjusted ORs increased with severity (P for trend <0.001): 1.23 (95% CI, 1.19–1.27) for mild, 2.36 (95% CI, 2.24–2.50) for moderate, and 7.39 (95% CI, 6.34–8.62) for severe hypotension. Sensitivity for correct MAP classification decreased from 87.1% for normal MAP to 32.0%, 19.6%, and 10.2% for mild, moderate, and severe hypotension.

**Conclusions:** We found a significantly higher risk of Cuff BP inaccuracy in patients with intraoperative hypotension, supporting cautious interpretation of Cuff BP during intraoperative hypotension.

**NOVELTY AND RELEVANCE:** *What Is New?:*

- Intraoperative hypotension was associated with increased inaccuracy of noninvasive oscillometric cuff blood pressure (Cuff BP) measurements. Inaccuracy increased progressively with hypotension severity, exceeding sevenfold greater odds during severe hypotension compared to normotension.

*What Is Relevant?:*

- Cuff BP is widely used for intraoperative monitoring, yet its accuracy under hypotensive conditions remains insufficiently established despite the importance of timely hypotension detection.

*Clinical/Pathophysiological Implications?:*

- Caution is warranted when interpreting Cuff BP during intraoperative hypotension. As hypotension becomes more severe and intervention more urgent, Cuff BP inaccuracy nevertheless increases.
- These findings have important perioperative patient-safety implications and support device validation under hypotensive and hemodynamically unstable intraoperative conditions.

## Introduction

Over 313 million major surgical procedures are performed annually worldwide and over 42 million surgeries are undertaken annually in the USA^1,2^. During surgeries, every patient receiving anesthesia has blood pressure (BP) monitoring at least every five minutes according to the American Society of Anesthesiologists^3^. For most patients, noninvasive oscilliometric cuff-based blood pressure (Cuff BP) measurements are widely utilized to monitor BP for close hemodynamic management during surgeries.

Intraoperative hypotension is one of the most common hemodynamic instabilities during surgery and is significantly associated with complications including postoperative myocardial infarction, acute kidney injury, and mortality^4–11^. It has been reported that over 90% of patients have at least one episode of blood pressure more than 20% below baseline^12^. Therefore, a high level of accuracy of BP measurement during surgery is crucial for the timely detection of intraoperative hypotension and prompt therapeutic intervention to prevent complications.

Despite the importance of high accuracy of Cuff BP, previous studies reported inaccuracy of Cuff BP^13–18^ due to diverse factors, including device- or patient-related sources^19^. Furthermore, the inaccuracy of Cuff BP in patients who have hypotension during surgery or in critically ill status was reported^20–23^. It may be because most Cuff BP devices were developed and validated in healthy study populations who usually have a normotensive baseline BP^21,24^. However, there is no large study investigating the inaccuracy of Cuff BP according to the level of hypotension during surgery.

To address this gap, we investigated the risk of Cuff BP inaccuracy in patients with intraoperative hypotension compared to those with intraoperative normotension among adult patients who underwent noncardiac surgery. Using Invasive arterial catheter-based blood pressure (Arterial BP) measurements as the gold standard BP measurement based on the previous study^13,22,25–27^, we analyzed a large U.S. intraoperative dataset containing numerous simultaneously measured pairs of Cuff BP and Arterial BP, along with potential risk factors for Cuff BP inaccuracy, demographic information, and comorbidities in patients undergoing diverse surgeries.

## Methods

### Study Population

We included adult patients who underwent noncardiac surgery at University of California, Los Angeles (UCLA) Medical Center from April 2013 to November 2023^28^. The UCLA perioperative data warehouse (PDW) has been described extensively in previous literature^28^.

Briefly, the UCLA PDW is a large-scale intraoperative dataset from electronic medical records (Epic Corporation) that includes patient demographic information, baseline co-morbidities, and operative procedure information. For this study, we excluded patients below age 18 years old according to the previous literature regarding the validation of BP measurement^16^. We also excluded those who underwent cardiac surgeries and those without simultaneous Cuff BP and Arterial BP measurements during surgery. We further excluded pairs measured outside of operating rooms to analyze only the pairs measured under anesthesiologists’ monitoring.

Because our dataset provides Arterial BP measurement locations (non-femoral [98.8%] vs. femoral [0.2%]), we excluded femoral readings to reduce heterogeneity arising from different anatomic sites and their effects on BP inaccuracy. This study was approved for Institutional Review Board (IRB) exception status by virtue of having no direct patient contact and using a deidentified dataset (IRB No. 15-000518).

### Blood Pressure Measurement and Monitoring

In accordance with institutional protocol, FDA-approved oscillometric Cuff BP and fluid-filled Arterial BP were utilized and calibrated at the heart level under the supervision of board-certified anesthesiologists. Detailed information regarding Cuff BP and Arterial BP measurement techniques, the devices used for Cuff BP and Arterial BP measurements, vital sign monitoring, and the algorithms employed to calculate MAP are provided in the Supplemental Methods.

### Exposure Ascertainment: Intraoperative Hypotension

To evaluate the association between intraoperative hypotension and Cuff BP inaccuracy, we used mean arterial pressure (MAP) to define intraoperative hypotension per previous literature^11^.We used the MAP values obtained from invasive arterial catheter-based blood pressure measurement, as this method is considered the gold standard for BP measurement during surgery^13,22,25–27^. To isolate the association between intraoperative hypotension and Cuff BP inaccuracy, analyses were restricted to normotensive and hypotensive BP ranges, excluding elevated BP measurements. We therefore excluded the pairs if systolic blood pressure (SBP) was higher than 120mmHg or diastolic blood pressure (DBP) was higher than 80mmHg, which are defined as elevated BP according to 2017 American Heart Association (AHA) high blood pressure guidelines^29^. According to the previous literature, we then defined intraoperative hypotension if MAP is less than 65mmHg and intraoperative normal BP if MAP is greater than 65^30–34^. To investigate the Cuff BP inaccuracy according to the severity of intraoperative hypotension, we further categorized MAP levels into four BP groups in an incremental manner by 10mmHg; severe hypotension (35 to <45mmHg), moderate hypotension (45 to <55mmHg), mild hypotension (55 to <65mmHg) and normal BP (≥ 65mmHg). We subsequently excluded instances of MAP below 35 mmHg, as these were considered outliers.

### Outcome Ascertainment: Cuff Blood Pressure Inaccuracy

For the primary outcome, we defined Cuff BP inaccuracy if the absolute difference between patient’s MAP measurement on Cuff BP and patient’s MAP measurement on Arterial BP is greater than 10 mmHg according to the previous statement of BP measuring device validation^16^. We used Arterial BP, the gold standard of BP measurement, as the reference BP to evaluate Cuff BP inaccuracy^13,22,25–27^. If the absolute difference of MAP of Cuff BP and Arterial BP is greater than 30mmHg, the pair was considered as outliers and excluded in the analysis. For a sensitivity analysis, we defined a modified outcome for intraoperative Cuff BP inaccuracy as a relative difference greater than 10% between MAP measured by Cuff BP and MAP measured by Arterial BP, accounting for the level of BP. We utilized a relative difference instead of an absolute difference for the modified outcome because, for high BP, an absolute difference of 10 mmHg may have a smaller effect, while for low BP, the same absolute difference may have a greater effect. This approach ensures that the measure is reflective of the BP context.

### Statistical Analysis

We analyzed the association between intraoperative hypotension and Cuff BP inaccuracy. Logistic regression models stratified by age (18 to <40, 40 to <50, 50 to <60, 60 to <70, and ≥70 years old) were used to calculate unadjusted and multivariable adjusted odds ratios (ORs) and 95% confidence intervals (CIs) of Cuff BP inaccuracy. Covariates were selected a priori based on putative risk factors and included sex (male, female), race (white, Black, Asian, other race), BMI (underweight, healthy, overweight, obesity), history of smoking, history of alcohol intake, hypertension, and diabetes (each yes/no), American Society of Anesthesiologists Physical Status (ASA PS) Classification (1-6), the type of surgery group (neurosurgery, general surgery, thoracic surgery, liver transplant, others), and intraoperative vasopressor use (no use, bolus use only, infusion use only, both bolus and infusion use). Further details regarding covariate categorization are provided in the Supplemental Methods.

For Model 1, it is an univariable model stratified by age without any additional adjustments. For multivariable model 2, we adjusted for sex, race, BMI, history of smoking, history of alcohol intake, hypertension, diabetes, ASA PS classification, and the type of surgery group. For multivariable model 3, we further adjusted for intraoperative vasopressor use. Missing data for categorical variables was included as a missing indicator. To evaluate the association between intraoperative hypotension and Cuff BP inaccuracy according to the severity of hypotension, we used the classified intraoperative hypotension as severe hypotension (35 to <45mmHg), moderate hypotension (45 to <55mmHg), and mild hypotension (55 to <65mmHg). Trend tests used the median of each category as a continuous variable.

For our primary analyses, we randomly selected 10 pairs of Cuff BP and Arterial BP per patient to ensure equal weighting across all patients. To select random pairs, we used a random selection algorithm in Python 3.12.2. If a patient had fewer than 10 pairs, all available pairs were included. To strengthen our findings, four additional analyses (classification performance, subgroup, exploratory, and sensitivity analyses) were conducted, with detailed statistical methods provided in the Supplemental Methods.

Two-sided p-values <0.05 were considered statistically significant. All analyses were performed using Python software version 3.12.2 (Python Software Foundation).

## Results

### Characteristics of the Study Population

We included 22,812 patients who have simultaneous measurements of Cuff BP and Arterial BP and 159,782 pairs of Cuff BP and Arterial BP from April 2013 to November 2023. Among patients having intraoperative hypotension (MAP < 65mmHg) on Arterial BP, mean (standard deviation; SD) of Arterial BP was 58.8 (5.3) mmHg and mean (SD) of Cuff BP was 65.7 (8.9) mmHg. Compared to patients having normal BP (MAP ≥ 65mmHg), patients having intraoperative hypotension (MAP < 65mmHg) were older, more likely to have history of smoking, more likely to have diabetes, and more likely to have intraoperative pressor use (**Table 1**).

**Table 1.**
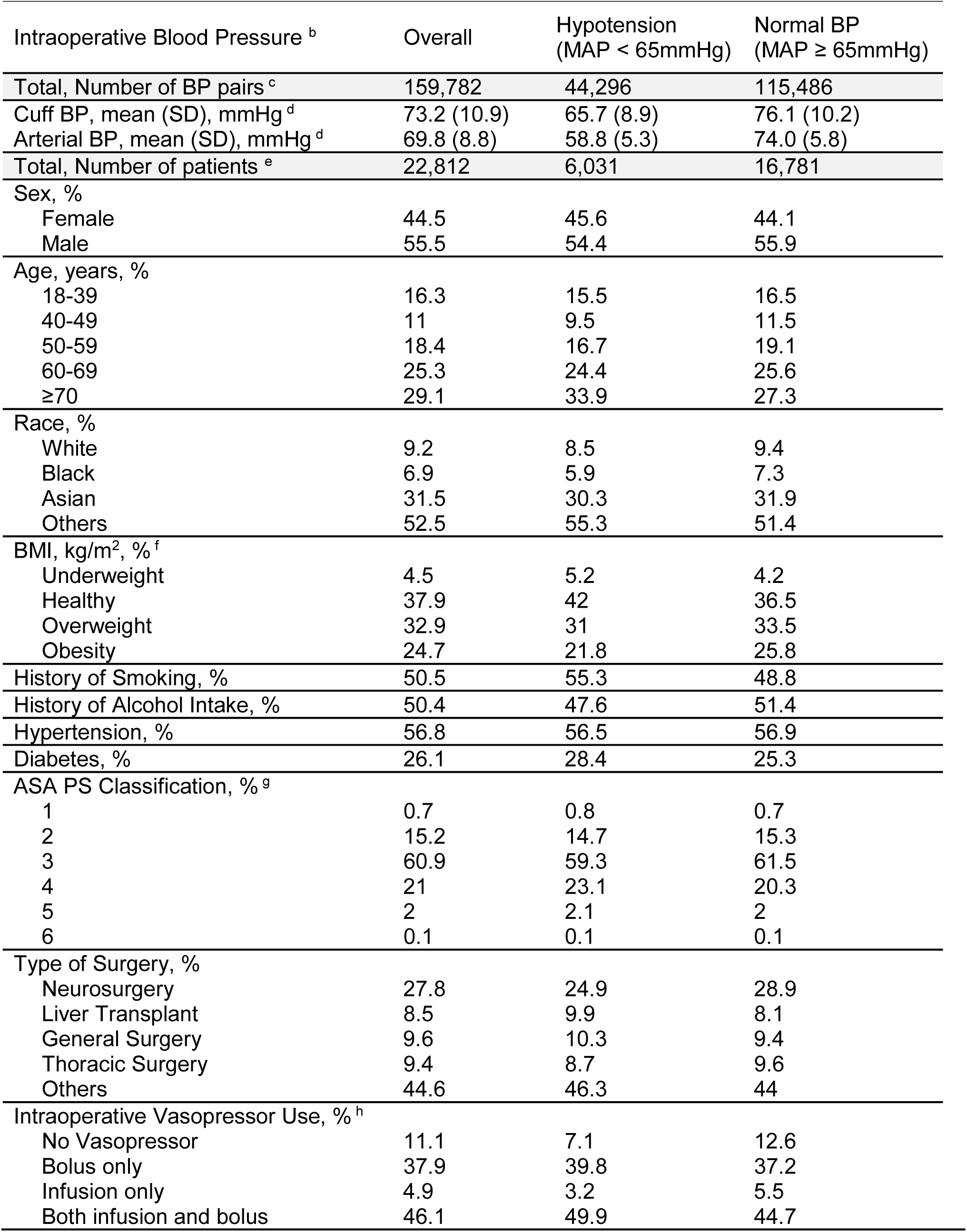
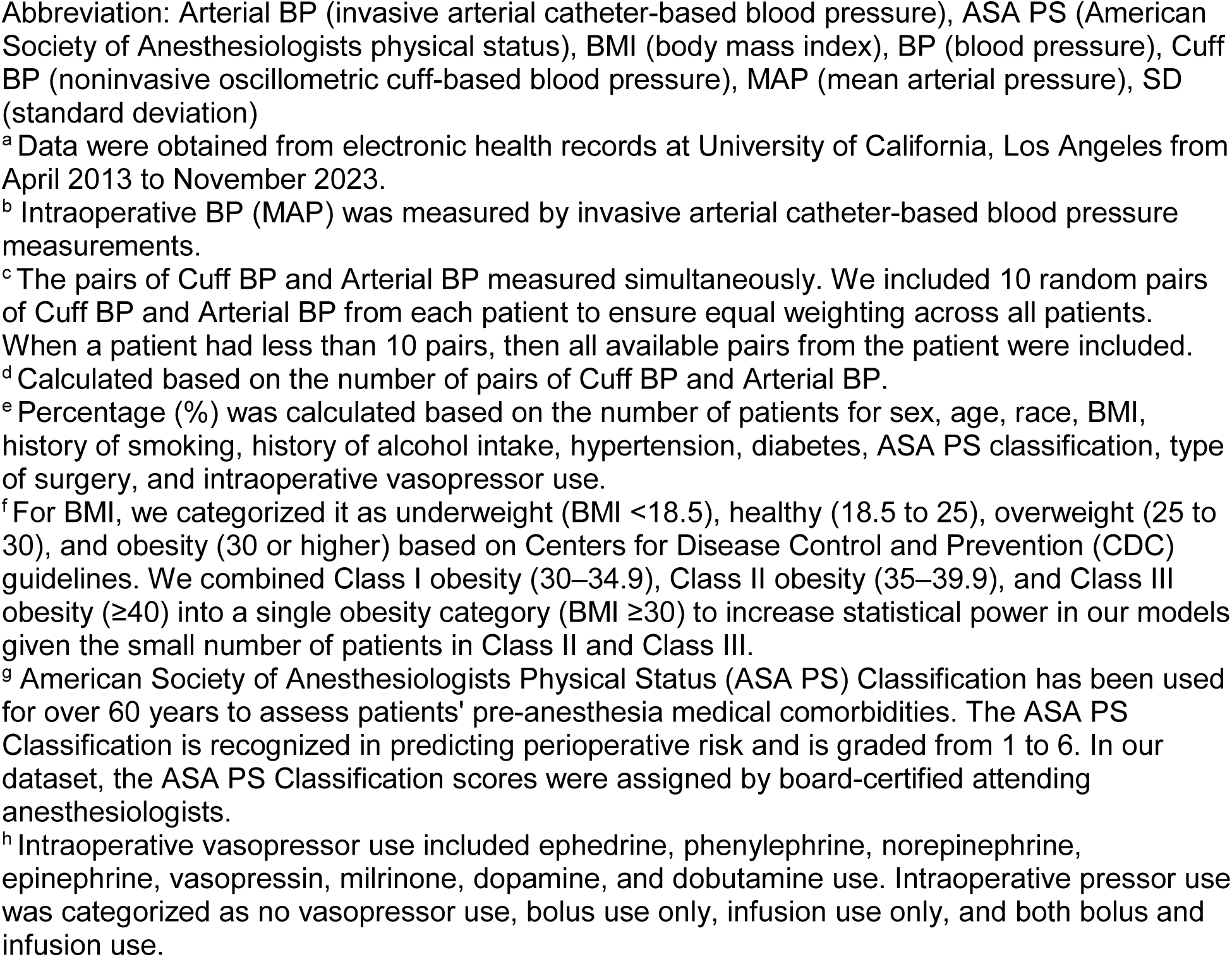
Baseline Characteristics of Study Population according to Intraoperative Blood Pressure ^a^.

| Intraoperative Blood Pressure <sup>b</sup> | Overall | Hypotension<br>(MAP < 65mmHg) | Normal BP<br>(MAP ≥ 65mmHg) |
| --- | --- | --- | --- |
| Total, Number of BP pairs <sup>c</sup> | 159,782 | 44,296 | 115,486 |
| Cuff BP, mean (SD), mmHg <sup>d</sup> | 73.2 (10.9) | 65.7 (8.9) | 76.1 (10.2) |
| Arterial BP, mean (SD), mmHg <sup>d</sup> | 69.8 (8.8) | 58.8 (5.3) | 74.0 (5.8) |
| Total, Number of patients <sup>e</sup> | 22,812 | 6,031 | 16,781 |
| Sex, % |  |  |  |
| Female | 44.5 | 45.6 | 44.1 |
| Male | 55.5 | 54.4 | 55.9 |
| Age, years, % |  |  |  |
| 18-39 | 16.3 | 15.5 | 16.5 |
| 40-49 | 11 | 9.5 | 11.5 |
| 50-59 | 18.4 | 16.7 | 19.1 |
| 60-69 | 25.3 | 24.4 | 25.6 |
| ≥70 | 29.1 | 33.9 | 27.3 |
| Race, % |  |  |  |
| White | 9.2 | 8.5 | 9.4 |
| Black | 6.9 | 5.9 | 7.3 |
| Asian | 31.5 | 30.3 | 31.9 |
| Others | 52.5 | 55.3 | 51.4 |
| BMI, kg/m<sup>2</sup>, % <sup>f</sup> |  |  |  |
| Underweight | 4.5 | 5.2 | 4.2 |
| Healthy | 37.9 | 42 | 36.5 |
| Overweight | 32.9 | 31 | 33.5 |
| Obesity | 24.7 | 21.8 | 25.8 |
| History of Smoking, % | 50.5 | 55.3 | 48.8 |
| History of Alcohol Intake, % | 50.4 | 47.6 | 51.4 |
| Hypertension, % | 56.8 | 56.5 | 56.9 |
| Diabetes, % | 26.1 | 28.4 | 25.3 |
| ASA PS Classification, % <sup>g</sup> |  |  |  |
| 1 | 0.7 | 0.8 | 0.7 |
| 2 | 15.2 | 14.7 | 15.3 |
| 3 | 60.9 | 59.3 | 61.5 |
| 4 | 21 | 23.1 | 20.3 |
| 5 | 2 | 2.1 | 2 |
| 6 | 0.1 | 0.1 | 0.1 |
| Type of Surgery, % |  |  |  |
| Neurosurgery | 27.8 | 24.9 | 28.9 |
| Liver Transplant | 8.5 | 9.9 | 8.1 |
| General Surgery | 9.6 | 10.3 | 9.4 |
| Thoracic Surgery | 9.4 | 8.7 | 9.6 |
| Others | 44.6 | 46.3 | 44 |
| Intraoperative Vasopressor Use, % <sup>h</sup> |  |  |  |
| No Vasopressor | 11.1 | 7.1 | 12.6 |
| Bolus only | 37.9 | 39.8 | 37.2 |
| Infusion only | 4.9 | 3.2 | 5.5 |
| Both infusion and bolus | 46.1 | 49.9 | 44.7 |

Abbreviation: Arterial BP (invasive arterial catheter-based blood pressure), ASA PS (American Society of Anesthesiologists physical status), BMI (body mass index), BP (blood pressure), Cuff BP (noninvasive oscillometric cuff-based blood pressure), MAP (mean arterial pressure), SD (standard deviation)
<sup>a</sup> Data were obtained from electronic health records at University of California, Los Angeles from April 2013 to November 2023.
<sup>b</sup> Intraoperative BP (MAP) was measured by invasive arterial catheter-based blood pressure measurements.
<sup>c</sup> The pairs of Cuff BP and Arterial BP measured simultaneously. We included 10 random pairs of Cuff BP and Arterial BP from each patient to ensure equal weighting across all patients. When a patient had less than 10 pairs, then all available pairs from the patient were included.
<sup>d</sup> Calculated based on the number of pairs of Cuff BP and Arterial BP.
<sup>e</sup> Percentage (%) was calculated based on the number of patients for sex, age, race, BMI, history of smoking, history of alcohol intake, hypertension, diabetes, ASA PS classification, type of surgery, and intraoperative vasopressor use.
<sup>f</sup> For BMI, we categorized it as underweight (BMI <18.5), healthy (18.5 to 25), overweight (25 to 30), and obesity (30 or higher) based on Centers for Disease Control and Prevention (CDC) guidelines. We combined Class I obesity (30–34.9), Class II obesity (35–39.9), and Class III obesity (≥40) into a single obesity category (BMI ≥30) to increase statistical power in our models given the small number of patients in Class II and Class III.
<sup>g</sup> American Society of Anesthesiologists Physical Status (ASA PS) Classification has been used for over 60 years to assess patients' pre-anesthesia medical comorbidities. The ASA PS Classification is recognized in predicting perioperative risk and is graded from 1 to 6. In our dataset, the ASA PS Classification scores were assigned by board-certified attending anesthesiologists.
<sup>h</sup> Intraoperative vasopressor use included ephedrine, phenylephrine, norepinephrine, epinephrine, vasopressin, milrinone, dopamine, and dobutamine use. Intraoperative pressor use was categorized as no vasopressor use, bolus use only, infusion use only, and both bolus and infusion use.

### Intraoperative Hypotension and Risk of Cuff BP Inaccuracy

For our primary analysis, we applied the primary outcome, Cuff BP inaccuracy, defined by absolute MAP difference between Cuff BP and Arterial BP greater than 10mmHg. Compared to having normal BP (MAP ≥ 65mmHg), having intraoperative hypotension (MAP < 65mmHg) was significantly associated with a higher risk of Cuff BP inaccuracy (Multivariable Model 2 OR 1.46, 95% CI 1.42-1.50; **Table 2**). The results remained almost unchanged after additional adjustment for intraoperative pressor use in Model 3. The risk of Cuff BP inaccuracy was substantially increased when severity of intraoperative hypotension was increased (P for trend < .001; **Table 3**). Compared to having normal BP (MAP ≥ 65mmHg), the multivariable ORs in Model 2 were 1.24 (95% CI 1.20-1.27) for mild hypotension (55 to < 65mmHg), 2.38 (95% CI 2.26-2.52) for moderate hypotension (45 to < 55mmHg), and 7.47 (95% CI 6.40-8.71) for severe hypotension (35 to < 45mmHg). The results were minimally changed after additional adjustment for intraoperative pressor use in Model 3.

**Table 2.** Risk of Cuff Blood Pressure Inaccuracy according to Intraoperative Blood Pressure ^a^.

|  | Intraoperative BP (MAP, mmHg) <sup>b</sup> |  |
| --- | --- | --- |
|  | Hypotension<br>(MAP < 65) | Normal BP<br>(65 ≤ MAP) |
| No. of Pairs <sup>c</sup> | 44,296 | 115,486 |
| No. of Cuff BP Inaccuracy <sup>d</sup> | 16,476 | 33,701 |
| Model 1 OR (95% CI) <sup>e</sup> | 1.43 (1.40-1.47) | 1 [Reference] |
| Model 2 OR (95% CI) <sup>f</sup> | 1.46 (1.42-1.50) | 1 [Reference] |
| Model 3 OR (95% CI) <sup>g</sup> | 1.45 (1.41-1.49) | 1 [Reference] |

Abbreviation: Arterial BP (invasive arterial catheter-based blood pressure), BP (blood pressure), CI (confidence interval), Cuff BP (noninvasive oscillometric cuff-based blood pressure), MAP (mean arterial pressure), OR (odds ratio)
Logistic regression models were used to calculate ORs and 95% CIs.
<sup>a</sup> Data were obtained from electronic health records at University of California, Los Angeles from April 2013 to November 2023.
<sup>b</sup> Mean arterial pressure was measured by invasive arterial catheter-based blood pressure measurements.
<sup>c</sup> The pairs of Cuff BP and Arterial BP measured simultaneously. We included 10 random pairs of Cuff BP and Arterial BP from each patient to ensure equal weighting across all patients. When a patient had less than 10 pairs, then all available pairs from the patient were included.
<sup>d</sup> Inaccuracy of Cuff BP was defined if absolute difference between Cuff BP and Arterial BP is greater than 10mmHg.
<sup>e</sup> Model 1 was adjusted for age (18 to <40, 40 to <50, 50 to <60, 60 to <70, and ≥70 years old).
<sup>f</sup> Model 2 was further adjusted for sex (male, female), race (white, Black, Asian, other race), body mass index (underweight, healthy, overweight, obesity), history of smoking, history of alcohol intake, hypertension, and diabetes (each yes/no), American Society of Anesthesiologists Physical Status Classification (1-6), and the type of surgery group (neurosurgery, general surgery, thoracic surgery, liver transplant, others).
<sup>g</sup> Model 3 was further adjusted for intraoperative vasopressor use (no use, bolus use only, infusion use only, both bolus and infusion use).

**Table 3.** Risk of Cuff Blood Pressure Inaccuracy according to Severity of Intraoperative Hypotension ^a^.

|  | Intraoperative BP (MAP, mmHg)<sup>b</sup> |  |  |  | P Value for Trends<sup>h</sup> |
| --- | --- | --- | --- | --- | --- |
|  | Severe Hypotension (35 ≤ MAP < 45) | Moderate Hypotension (45 ≤ MAP < 55) | Mild Hypotension (55 ≤ MAP < 65) | Normal BP (65 ≤ MAP) |  |
| No. of Pairs<sup>c</sup> | 1,152 | 7,437 | 35,707 | 115,486 |  |
| No. of Cuff BP Inaccuracy<sup>d</sup> | 832 | 3,602 | 12,042 | 33,701 |  |
| Model 1 OR (95% CI)<sup>e</sup> | 6.27 (5.51-7.14) | 2.27 (2.16-2.38) | 1.23 (1.20-1.26) | 1 [Reference] | <.001 |
| Model 2 OR (95% CI)<sup>f</sup> | 7.47 (6.40-8.71) | 2.38 (2.26-2.52) | 1.24 (1.20-1.27) | 1 [Reference] | <.001 |
| Model 3 OR (95% CI)<sup>g</sup> | 7.39 (6.34-8.62) | 2.36 (2.24-2.50) | 1.23 (1.19-1.27) | 1 [Reference] | <.001 |

Abbreviation: Arterial BP (invasive arterial catheter-based blood pressure), BP (blood pressure), CI (confidence interval), Cuff BP (noninvasive oscillometric cuff-based blood pressure), MAP (mean arterial pressure), OR (odds ratio)
Logistic regression models were used to calculate ORs and 95% CIs.
<sup>b</sup> Mean arterial pressure was measured by invasive arterial catheter-based blood pressure measurements.
<sup>c</sup> The pairs of Cuff BP and Arterial BP measured simultaneously. We included 10 random pairs of Cuff BP and Arterial BP from each patient to ensure equal weighting across all patients. When a patient had less than 10 pairs, then all available pairs from the patient were included.
<sup>d</sup> Inaccuracy of Cuff BP was defined if absolute difference between Cuff BP and Arterial BP is greater than 10mmHg.
<sup>e</sup> Model 1 was adjusted for age (18 to <40, 40 to <50, 50 to <60, 60 to <70, and ≥70 years old).
<sup>f</sup> Model 2 was further adjusted for sex (male, female), race (white, Black, Asian, other race), body mass index (underweight, healthy, overweight, obesity), history of smoking, history of alcohol intake, hypertension, and diabetes (each yes/no), American Society of Anesthesiologists Physical Status Classification (1-6), and the type of surgery group (neurosurgery, general surgery, thoracic surgery, liver transplant, others).
<sup>g</sup> Model 3 was further adjusted for intraoperative vasopressor use (no use, bolus use only, infusion use only, both bolus and infusion use).
<sup>h</sup> Two-sided P values for trend were calculated using the median value of each category as a continuous variable.

### Classification Performance of Cuff BP by Severity of Intraoperative Hypotension

To evaluate the Cuff BP classification performance according to the severity of intraoperative hypotension, we applied the confusion matrix analysis and calculated sensitivity and positive predictive value (PPV). Sensitivity was 87.1% for normal BP (MAP ≥ 65mmHg), but substantially decreased to 32.0% for mild hypotension (55 to < 65mmHg), 19.6% for moderate hypotension (45 to < 55mmHg), and 10.2% for severe hypotension (35 to < 45mmHg) (**Figure 2**). Similarly, PPV was 80.7% for normal BP (MAP ≥ 65mmHg), but declined markedly to 41.0% for mild hypotension (55 to < 65mmHg), 22.5% for moderate hypotension (45 to < 55mmHg), and 14.4% for severe hypotension (35 to < 45mmHg). In addition, to evaluate the difference between Cuff BP and Arterial BP across intraoperative BP levels, we applied a Bland-Altman plot. The plot demonstrated that the mean and SD of the differences between Cuff BP and Arterial BP increased noticeably as intraoperative BP decreased (**Supplemental Figure 1**).

**Figure 1.**
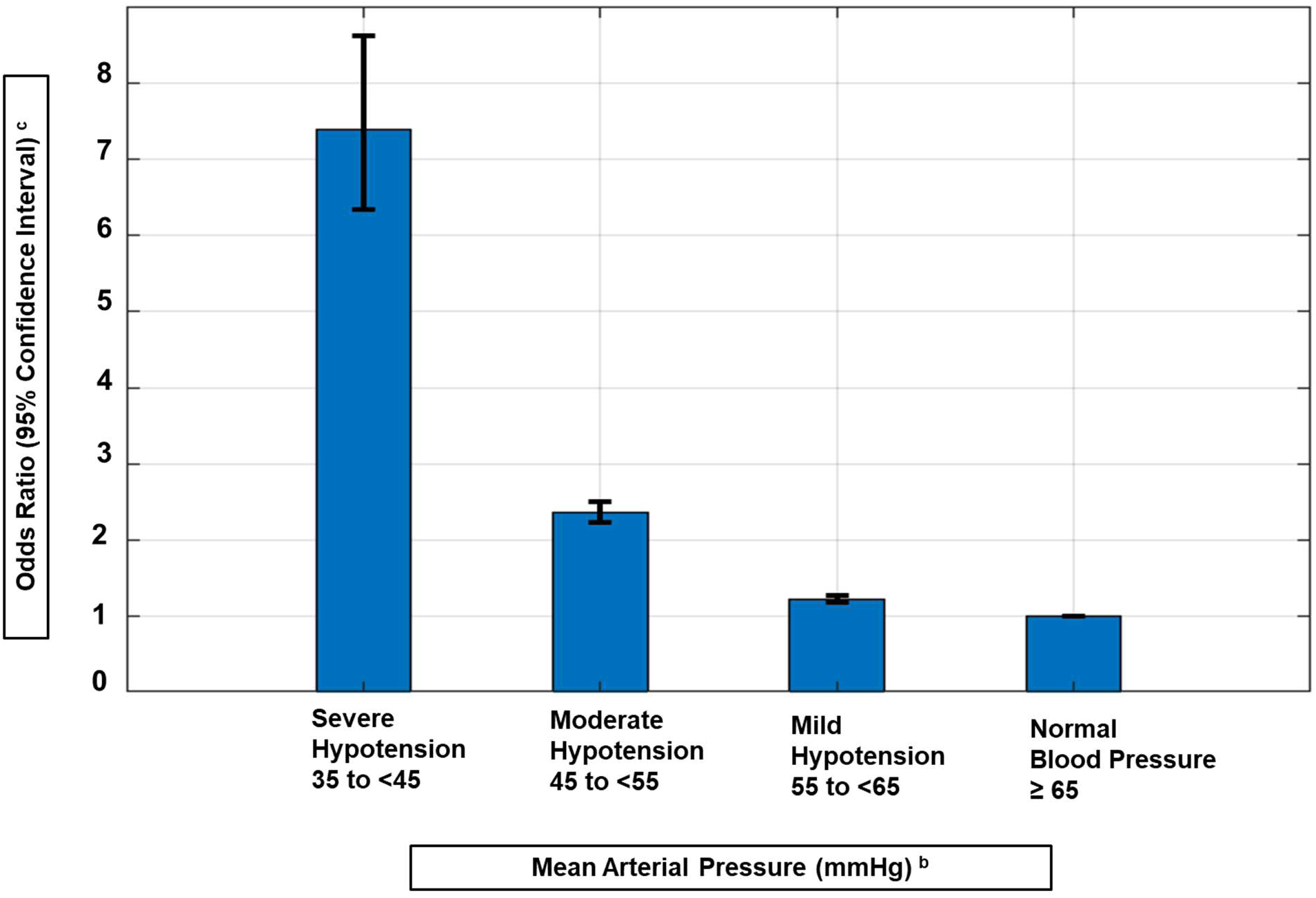
Risk of Cuff Blood Pressure Inaccuracy according to Severity of Intraoperative Hypotension ^a^. ^a^ Data were obtained from electronic health records at University of California, Los Angeles from April 2013 to November 2023. ^b^ Mean arterial pressure was measured by invasive arterial catheter-based blood pressure measurements. ^c^ Odds Ratio and 95% Confidence Interval were calculated using Model 3. Model 3 was adjusted for age (18 to <40, 40 to <50, 50 to <60, 60 to <70, and ≥70 years old), sex (male, female), race (white, Black, Asian, other race), body mass index (underweight, healthy, overweight, obesity), history of smoking, history of alcohol intake, hypertension, and diabetes (each yes/no), American Society of Anesthesiologists Physical Status Classification (1-6), and the type of surgery group (neurosurgery, general surgery, thoracic surgery, liver transplant, others), intraoperative vasopressor use (no use, bolus use only, infusion use only, both bolus and infusion use).

**Figure 2.**
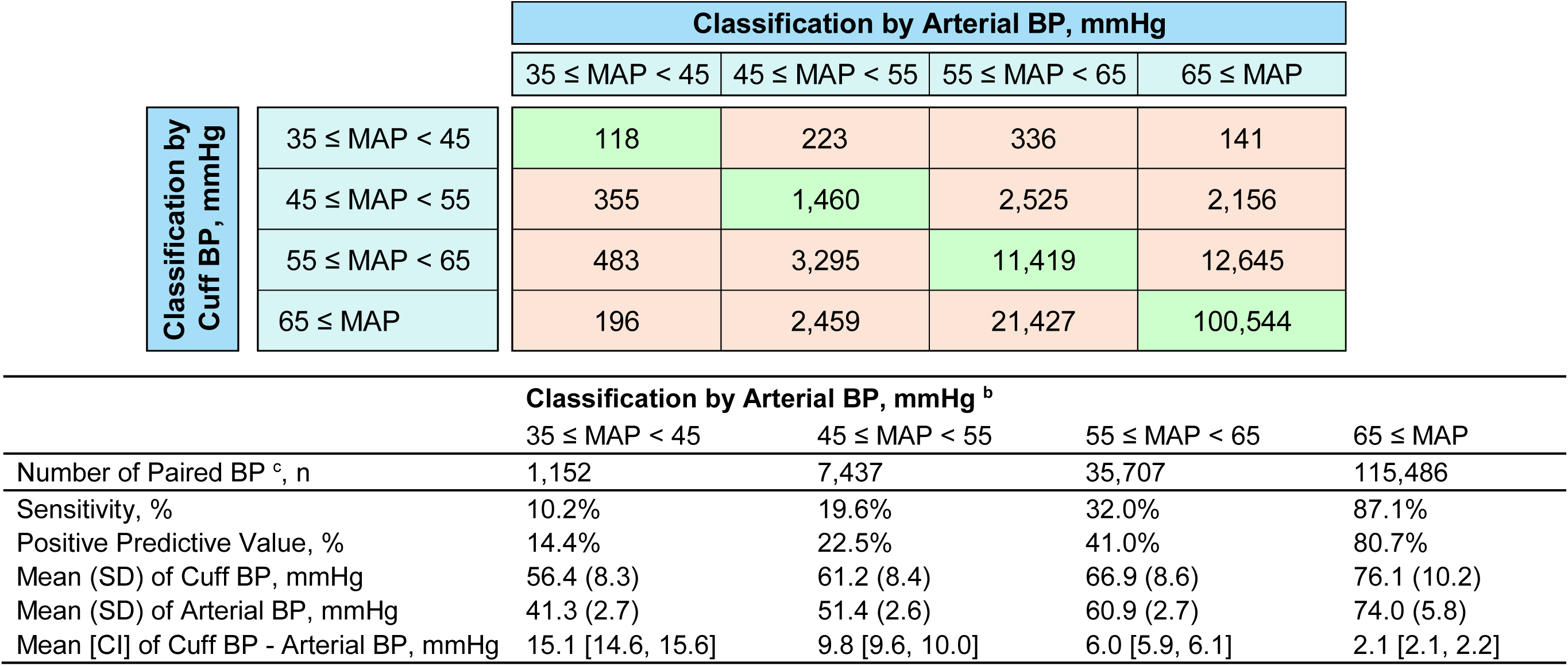
Classification Performance of Cuff Blood Pressure Measurements according to Severity of Intraoperative Hypotension ^a^. Abbreviation: Arterial BP (invasive arterial catheter-based blood pressure), BP (blood pressure), Cuff BP (noninvasive oscillometric cuff-based blood pressure), MAP (mean arterial pressure), SD (standard deviation) ^a^ Data were obtained from electronic health records at University of California, Los Angeles from April 2013 to November 2023. ^b^ Mean arterial pressure was measured by invasive arterial catheter-based blood pressure measurements. ^c^ The pairs of Cuff BP and Arterial BP measured simultaneously. We included 10 random pairs of Cuff BP and Arterial BP from each patient to ensure equal weighting across all patients. When a patient had less than 10 pairs, then all available pairs from the patient were included.

### Subgroup and Stratified Analyses

In subgroup analysis, we assessed for effect modification by demographic factors including sex, age, BMI, and comorbidities. We observed an association between intraoperative hypotension and increased risk of Cuff BP inaccuracy were more pronounced among males, individuals older than 65, individuals with a history of hypertension, and individuals with a history of diabetes (**Supplemental Table 1**). In stratified analysis in each surgery group, the association between intraoperative hypotension and the increased risk of Cuff BP inaccuracy were consistent with statistical significance within each surgery group (**Supplemental Table 2**). Compared to having normal BP (MAP ≥ 65mmHg), the multivariable ORs for risk of Cuff BP inaccuracy were 1.37 (95% CI 1.30-1.44) for neurosurgery, 1.58, (95% CI 1.44-1.73) for general surgery, 1.24 (95% CI 1.13-1.36) for thoracic surgery, 1.58 (95% CI 1.44-1.74) for liver transplant, 1.59 (95% CI 1.42-1.77) for orthopedics, 1.51 (95% CI 1.35-1.69) for urology, 1.55 (95% CI 1.41-1.71) for otolaryngology, 1.54 (95% CI 1.37-1.74) for vascular surgery, 1.43 (95% CI 1.27-1.60) for surgical oncology, and 1.71 (95% CI 1.38-2.13) for obstetrics and gynecology.

### Exploratory and Sensitivity Analyses

In an exploratory analysis, we used averaged BP from three paired BP samples measured sequentially within a 5-minute interval, in accordance with the AHA recommendation to obtain two to three consecutive BP measurements for a more accurate assessment^35^. The results were consistent with our primary results (multivariable OR 1.43, 95% CI 1.28-1.59; **Supplemental Table 3**). For sensitivity analysis, we first investigated the association using a modified outcome, defined as Cuff BP inaccuracy when the relative MAP difference between Cuff BP and Arterial BP is greater than 10%. The results were consistent with our primary results (multivariable OR 1.97, 95% CI 1.92-2.03; **Supplemental Table 5**). Additionally, further adjustment for the history of ischemic heart disease, congestive heart failure, or ASA emergency status did not substantially alter the association (multivariable OR, 1.45; 95% CI, 1.41-1.49).

## Discussion

In this large size perioperative cohort study of 22,812 patients undergoing noncardiac surgery at a single quaternary referral center in the U.S, we observed that intraoperative hypotension was associated with a 45% higher risk of Cuff BP inaccuracy compared to normal BP. Notably, the increased risk of Cuff BP inaccuracy was even more pronounced among patients with moderate hypotension and reached a maximum among those with severe hypotension, exhibiting a more than 7-fold higher risk.

Our findings are consistent with the previous studies investigating inaccuracy of Cuff BP when patient is hypotensive^20–23,30^. A prospective observational study in emergency department found that MAP on oscillometric device using an upper arm cuff was markedly higher than MAP on arterial catheter among patients having hypotension (MAP <60mmHg)^21^. Similarly, a study using electronic anesthesia records observed that noninvasive BP was generally higher than intra-arterial BP during periods of hypotension^20^. The other study using a large ICU database also found that clinically significant discrepancies between invasive arterial BP and oscillometric cuff BP measurement during hypotension^36^. Additionally, a study including 85 patients hospitalized for cardiogenic shock found that clinical conditions associated with hypotension generally worsened the accuracy of noninvasive MAP^23^. However, the previous studies did not evaluate the risk of Cuff BP inaccuracy according to the severity of hypotension or had an insufficient sample size of patients. Thus, our results significantly expand upon prior studies by examining the severity of hypotension using numerous pairs of Cuff BP and Arterial BP, particularly during surgery, when patients are especially susceptible to hypotension.

The mechanism of oscillometric cuff based BP is detecting arterial blood flow while inflating and deflating the cuff encircling a limb^37,38^. The inaccuracy of oscillometric cuff BP may be caused by multiple factors. Arterial stiffness from calcification or aging, an improperly sized cuff relative to the patient’s arm circumference, arm shape, and irregular pulses can all contribute to inaccuracy in Cuff BP measurement^19,30,37–39^. More importantly, because Cuff BP is calibrated by manufacturers using a healthy population with typically normotensive BP^21,24,38^, it may be inaccurate for patients whose BP falls substantially outside the normotensive range. In particular, Cuff BP among patients with hypotension and shock is likely to be inaccurate due to abnormal vascular tone or low cardiac output compared with healthy population. Finally, the lack of standardized Cuff BP validation for hemodynamically unstable patients may lead to significant measurement inaccuracy.

High accuracy of BP measurement during surgery is critically needed for timely therapeutic intervention because intraoperative hypotension is highly associated with perioperative complications^7–10,40–44^. A prior literature reported that intraoperative hypotension with MAP below absolute thresholds of 65 mmHg or relative thresholds of 20% were related to both myocardial and kidney injury^43^. Furthermore, a previous retrospective cohort study demonstrated that intraoperative hypotension is associated with increased 30-day operative mortality in patients having noncardiac surgery^7,44^. A substudy of randomized trial also reported that intraoperative hypotension was associated with a composite of myocardial infarction and death during perioperative periods^8^. Therefore, accurate BP measurement during surgery is critically needed because intraoperative hypotension is potentially modifiable to decrease perioperative adverse outcomes.

Our study demonstrated a remarkably apparent association between intraoperative hypotension and Cuff BP inaccuracy among patients having moderate hypotension, with the association being most pronounced among patients with severe hypotension. Additionally, in an exploratory analysis, we observed findings consistent with our primary results when using averaged BP obtained from three sequential Cuff BP measurements, in accordance with the AHA recommendation to obtain two to three consecutive BP measurements for a more accurate assessment^35^. Notably, on sensitivity analyses, when we redefined Cuff BP inaccuracy as a relative MAP difference of greater than 10% instead of an absolute MAP difference of 10 mmHg, we observed that the association became more pronounced, demonstrating relative MAP difference is more reflective of the BP context.

In the subgroup analysis, we observed that the association between intraoperative hypotension and the increased risk of Cuff BP inaccuracy was more pronounced among males, individuals aged over 65, individuals with a history of hypertension, and individuals with a history of diabetes. Individuals with advanced age, hypertension, and diabetes are more susceptible to vascular changes which may further contribute to Cuff BP inaccuracy.

### Strengths and Limitations

This study has several strengths. First, this study utilized a large intraoperative cohort that included numerous concurrent measurements of Cuff BP and Arterial BP. This unique dataset enabled a comprehensive investigation of Cuff BP inaccuracy among patients with intraoperative hypotension. Second, this study investigates the risk of Cuff BP inaccuracy according to the extent of hypotension. Although hypotension has been recognized as a contributor to Cuff BP inaccuracy, the association between the severity of intraoperative hypotension and Cuff BP inaccuracy has not been well characterized. Third, this study robustly adjusts for potential confounders, including types of surgery and intraoperative vasopressor use, as well as demographic characteristics and comorbidities. Fourth, this study utilizes a high-fidelity dataset with minimal measurement error. This large dataset was collected intraoperatively and automatically through electronic medical records, minimizing human errors in BP measurement charting. Furthermore, BP monitoring devices were placed, calibrated, and closely monitored by board-certified anesthesiologists according to ASA protocols, ensuring high-quality data. Last, the study population underwent a wide variety of surgical procedures because the cohort was derived from a quaternary referral center. This enabled investigation of the association between intraoperative hypotension and Cuff BP inaccuracy across diverse surgical populations.

This study has several limitations. First, although we included a wide range of risk factors relevant to Cuff BP inaccuracy, we cannot rule out the potential for residual confounding since our study is observational. Second, Arterial BP is also subject to measurement error from several sources, including arterial line quality and patient-specific vascular factors. Although these sources of error cannot be completely eliminated, their impact was likely minimized in our study. In this study, Arterial BP monitoring was overseen by board-certified anesthesiologists at a quaternary academic referral center, where arterial line quality was periodically verified according to institutional practice. Furthermore, our multivariable models adjusted for factors that may influence Arterial BP measurements, including hypertension, diabetes mellitus, and intraoperative vasopressor use. Therefore, consistent with previous studies^13,22,25–27^, we accepted Arterial BP as the reference standard for intraoperative BP measurement. Third, these data do not include information about the size of the cuff used for Cuff BP reading. However, the placement and measurement of Cuff BP were performed and confirmed under the supervision of board-certified anesthesiologists, which minimized the effect on Cuff BP inaccuracy due to an inappropriate cuff size. Fourth, this dataset does not have information about the location of BP measurement. However, previous studies have demonstrated that MAP remains minimally changed between the ascending aorta and peripheral artery, whereas systolic pressure varies depending on the body location^45^. Therefore, we used MAP in our main analyses to minimize variability in BP caused by different measurement locations. Ideally, Cuff BP and Arterial BP would be measured simultaneously at the same anatomical location to eliminate any potential errors. However, this is not feasible because the arterial line signal disappears while the cuff is inflating (Supplemental Discussion). Fifth, this dataset did not include information on patient positioning. However, the BP monitoring devices, including the transducer, were leveled at the same height as the patient’s heart during surgery by board-certified anesthesiologists. Sixth, we intentionally excluded BP measurements in the hypertensive range because intraoperative hypertension may arise from mechanisms distinct from those underlying hypotension, such as inadequate pain-control or sedation rather than reduced cardiac output or vascular tone. Future studies should specifically assess Cuff BP accuracy during intraoperative hypertension. Last, these data were derived from a single center and collected from patients who underwent noncardiac surgeries with anesthetic agents and invasive arterial catheter placement, which may reduce generalizability.

In conclusion, in this large intraoperative cohort, intraoperative hypotension was associated with an increased risk of Cuff BP inaccuracy, and this risk increased progressively with hypotension severity. These findings suggest that Cuff BP measurements should be interpreted cautiously during intraoperative hypotension.

## PERSPECTIVES

Cuff BP is used nearly universally for intraoperative BP monitoring, yet its accuracy has not been adequately validated under the hemodynamic conditions encountered during surgery. Our findings demonstrate that Cuff BP becomes progressively less reliable as intraoperative hypotension becomes more severe, precisely when accurate BP measurement is most important for clinical decision-making. These findings highlight the need for device-validation protocols that assess accuracy of BP monitoring devices across clinically relevant hypotensive ranges and in vulnerable surgical populations. Future studies should determine whether improved validation standards, repeated measurements, or alternative monitoring strategies can reduce Cuff BP inaccuracy and improve patient management during surgery, given the increasing number of surgical procedures, the universal use of Cuff BP intraoperatively, and the high incidence of intraoperative hypotension and its perioperative complications.

## Data Availability

The datasets generated and/or analyzed during the current study are not publicly available due to privacy and ethical restrictions, but are available from the corresponding author on reasonable request.

## Role of the Funder/Sponsor

Funders/Sponsors had no role in study design, analysis, and interpretation of data, report writing, and the decision to submit for publication.

The corresponding author had full access to data and the final responsibility to submit for publication.

## ABBREVIATION

**Abbreviation Definition**

AHA: American Heart Association
Arterial BP: invasive arterial catheter-based blood pressure
ASA PS: American Society of Anesthesiologists Physical Status
BMI: body mass index
BP: blood pressure
CDC: Centers for Disease Control and Prevention
CI: confidence interval
Cuff BP: noninvasive oscillometric cuff-based blood pressure
DBP: diastolic blood pressure
IRB: Institutional Review Board
MAP: mean arterial pressure
OR: odds ratio
PDW: perioperative data warehouse
PPV: positive predictive value
SBP: systolic blood pressure
SD: standard deviation

